# The early-stage comprehensive costs of routine PrEP implementation and scale-up in Zambia

**DOI:** 10.1101/2021.12.18.21268023

**Authors:** Cheryl Hendrickson, Lawrence C Long, Craig van Rensburg, Cassidy W Claassen, Mwansa Njelesani, Crispin Moyo, Lloyd Mulenga, Heidi O’Bra, Colin A Russell, Brooke E Nichols

## Abstract

**Introduction:** Pre-exposure prophylaxis (PrEP) is effective at preventing HIV infection, but PrEP cost-effectiveness is sensitive to PrEP implementation and program costs. Preliminary studies indicate that, in addition to direct delivery cost, PrEP provision requires substantial demand creation and user support to encourage PrEP initiation and persistence. We estimated the cost of providing PrEP in Zambia through different PrEP delivery models.

**Methods:** Taking a guidelines-based approach for visits, labs and drugs assuming fidelity to the expanded 2018 Zambian PrEP guidelines, we estimated the annual cost of providing PrEP per client for five delivery models: one focused on key populations (men-who-have-sex-with-men (MSM) and female sex workers (FSW), one on adolescent girls and young women (AGYW), and three integrated programs (operated within the HIV counselling and testing service at primary healthcare centres). Program start-up, provider, and user support costs were based on program expenditure data and number of PrEP sites and clients in 2018. PrEP clinic visit costs were based on micro-costing at two PrEP delivery sites (in 2018 USD).

**Results:** The annual cost per PrEP client varied greatly by program type, from $394 (AGYW) to $760 in an integrated program. Cost differences were driven largely by volume (i.e. the number of clients initiated/model/site) which impacted the relative costs of program support and technical assistance assigned to each PrEP client. Direct service delivery costs, including staff and overheads, labs and monitoring, drugs and consumables ranged narrowly from $208-217/PrEP-user. Service delivery costs were a key component in the cost of PrEP, representing 36-65% of total costs. Reductions in service delivery costs per PrEP client are expected with further scale-up.

**Conclusions:** The results show that, even when integrated into full service delivery models, accessing vulnerable, marginalised populations at substantial risk of HIV infection is likely to cost more than previously estimated due to the programmatic costs involved in community sensitization and user support. Improved data on individual client resource usage (e.g. drugs, labs, visits) and outcomes (e.g. initiation, persistence) is required to get a better understanding of the true resource utilization, cost and expected outcomes and annual costs of different PrEP programs in Zambia.

## Introduction

Following several clinical trials and observational studies (1-3), the World Health Organization (WHO) has recommended that pre-exposure prophylaxis (PrEP) be offered to anyone at substantial risk of HIV infection (4). Several low- and middle-income countries (LMICs) have adopted these guidelines and are currently implementing national PrEP programs (5). However, limited data are available to inform the total cost and affordability of these programs, which is crucial for policy decisions around implementation strategies for HIV prevention programs.

Several studies have modelled the cost effectiveness of PrEP provision with mixed results (6-23); however, they largely focus on direct service delivery costs and many do not take into account routine PrEP program support costs. The costs of PrEP programs implemented in routine care settings in sub-Saharan Africa (SSA) are largely unreported and those that are, have thus far focused primarily on targeting key populations including female sex workers (FSW), men who have sex with men (MSM), and priority populations such as adolescent girls and young women (AGYW). Evidence demonstrating the variations in costs of these different PrEP programs, service delivery strategies, and resource utilization in SSA is limited, with only a few demonstration projects having published the costs of providing PrEP in these settings (24-29).

Estimating the costs of scaling up PrEP programs is complicated by the changing ‘population-at-risk’ for any given time period. Unlike ART, which is intended as a life-long treatment program under which optimal treatment requires 100% adherence, PrEP is effective when taken during periods of high risk, meaning that cycling on and off PrEP may be appropriate (30). This complicates analyses of budgetary needs and scale-up costs of PrEP programs, an issue that Zambia is currently facing with national PrEP implementation and scale-up. Zambia, a lower-middle income country in southern Africa with an estimated 48,000 new HIV infections in 2018, is currently investing in scaling up its PrEP program (31). Zambia has adopted daily oral PrEP containing tenofovir and emtricitabine as an additional HIV prevention strategy, recommending PrEP for all HIV-negative persons at high risk of HIV acquisition (32). By September 2018, PrEP was offered in 162 sites across nine of the ten provinces, with 3,626 people at risk of HIV infection initiated on PrEP; one year later, this had increased to 23,327 at 728 sites across all provinces(33). These were largely serodiscordant couples, but also included AGYW ages 15-24 (35%), FSW (9%), and MSM (3%) (34). The Zambian Ministry of Health aims to further expand PrEP services, but has limited information on the cost and budgetary impact of this national scale-up.

We present the results of a costing study of PrEP implementation in Zambia, aiming to provide cost estimates of PrEP provision disaggregated by program type. We also present costs per PrEP-month of effective use, using aggregate PrEP persistence data, and compare that to costs for perfect use.

## Methods

### Study Setting

Zambia’s PrEP guidelines stipulate that those eligible for PrEP and identified as being at substantial risk of HIV infection (as defined as engaging in one or more risky activities in the past six months) be offered PrEP (Table 1). We costed three types of service delivery models that offered PrEP to: 1) anyone at risk of HIV infection visiting a primary healthcare facility. These programs had integrated PrEP into their HIV testing and counselling (HCT) services) (integrated; across three implementing partners); 2) AGYW as part of a broad, multi-component package of evidence-based health, educational and social interventions aimed at reducing new HIV infections (AGYW-focused); and 3) FSW and MSM through tailored, targeted community-based interventions (FSW/MSM-focused). The three integrated models, each led by different implementation partners, were rolled out differently and incurred slightly different program costs. The first model focused on outreach, using trained community health workers to sensitize the community about PrEP and refer those interested in PrEP to the local clinic. The second model focused resources on site readiness through preliminary site assessments, training and community consultations. Additionally, there were several international technical support visits from implementation partners. The third model was based on the Community HIV Epidemic Model (CHEC) of care (35), with costs incurred for community education, mobilization, and PrEP sensitization alongside training of health care workers (HCWs) on KP sensitivity and PrEP service delivery. The AGYW-focused program was integrated as part of the DREAMS program for adolescent girls and young women (36). DREAMS (Determined, Resilient, Empowered, AIDS-free, Mentored and Safe) partnership is a public-private partnership aimed at reducing rates of HIV among AGYW in highest HIV burden countries. It is largely funded and implemented by the United States Agency for International Development (USAID). Support services in this DREAMS PrEP-provision model included community sensitization and demand creation activities through short-term community mobilisers. A website focused on PrEP for AGYW was also created. PrEP was provided at specialized DREAMS centers that had an integrated community health worker who was trained on PrEP provision. The program also provided continued PrEP-user support using peer navigators. Finally, the FSW- and MSM-focused programs were targeted community-based demand creation programs with referrals to local facilities for PrEP initiation and follow up. Short-term mobilisers and peer navigators spent time in the local communities to specifically reach FSW and MSM populations, create demand for PrEP refer them for services at the clinics and provide continued support to PrEP clients. A three and a half-day non-clinical training was undertaken to orient non-clinical program staff on PrEP roll-out.

**Table 1:** PrEP clinical activities adapted from the Zambian PrEP implementation framework (38)

|  | Year 1 (at least 6 visits) |  |  |  |  |  | Year 2 (at least 4 visits) |  |  |  |
| --- | --- | --- | --- | --- | --- | --- | --- | --- | --- | --- |
|  | Initial visit and<br>PrEP initiation | Follow-up months |  |  |  |  | 15 | 18 | 21 | 24 |
|  |  | 1 | 3 | 6 | 9 | 12 |  |  |  |  |
| Confirmation of HIV-negative status | x | x | x | x | x | x | x | x | x | x |
| Serum Creatinine | x | x | x | x | x | x |  |  |  | x |
| Hepatitis B surface antigen | x |  |  | x |  |  |  |  |  |  |
| Hepatitis C antibody* | x |  |  | x |  |  |  |  |  |  |
| Pregnancy testing** | x |  |  |  |  |  |  |  |  |  |
| Rapid Plasma Reagin (RPR) | x |  |  | x |  |  |  | x |  |  |
| Other screening for sexually<br>transmitted infections (STIs) | x |  | x | x | x | x | x | x | x | x |
| Alanine Transaminase (ALT) | x |  |  | x |  |  |  |  |  | x |

\*Included for men-who-have-sex-with-men MSM only
\*\*Included for women only

### Costs

The costs of the PrEP programs were estimated from the providers’ perspective, with data collected in 2018 and reported in 2018 US dollars (USD). Costs collected in Zambian Kwacha were converted to the USD equivalent based on the average exchange rate for 2018 of 10.4988 ZMW to 1 USD (January-December) (37). We used expenditure analysis in estimating financial costs and tracked actual implementation expenses including training, demand creation and technical support. At the facility-level we only included costs directly related to PrEP programming such as counsellor-time, facility space and equipment. We categorized the costs into program costs and direct service delivery costs. Program costs included project start-up costs, training, sensitization as well as provider and user support costs. Direct service delivery costs included clinical personnel, supplies, laboratory tests and drugs (Table 2).

**Table 2:**
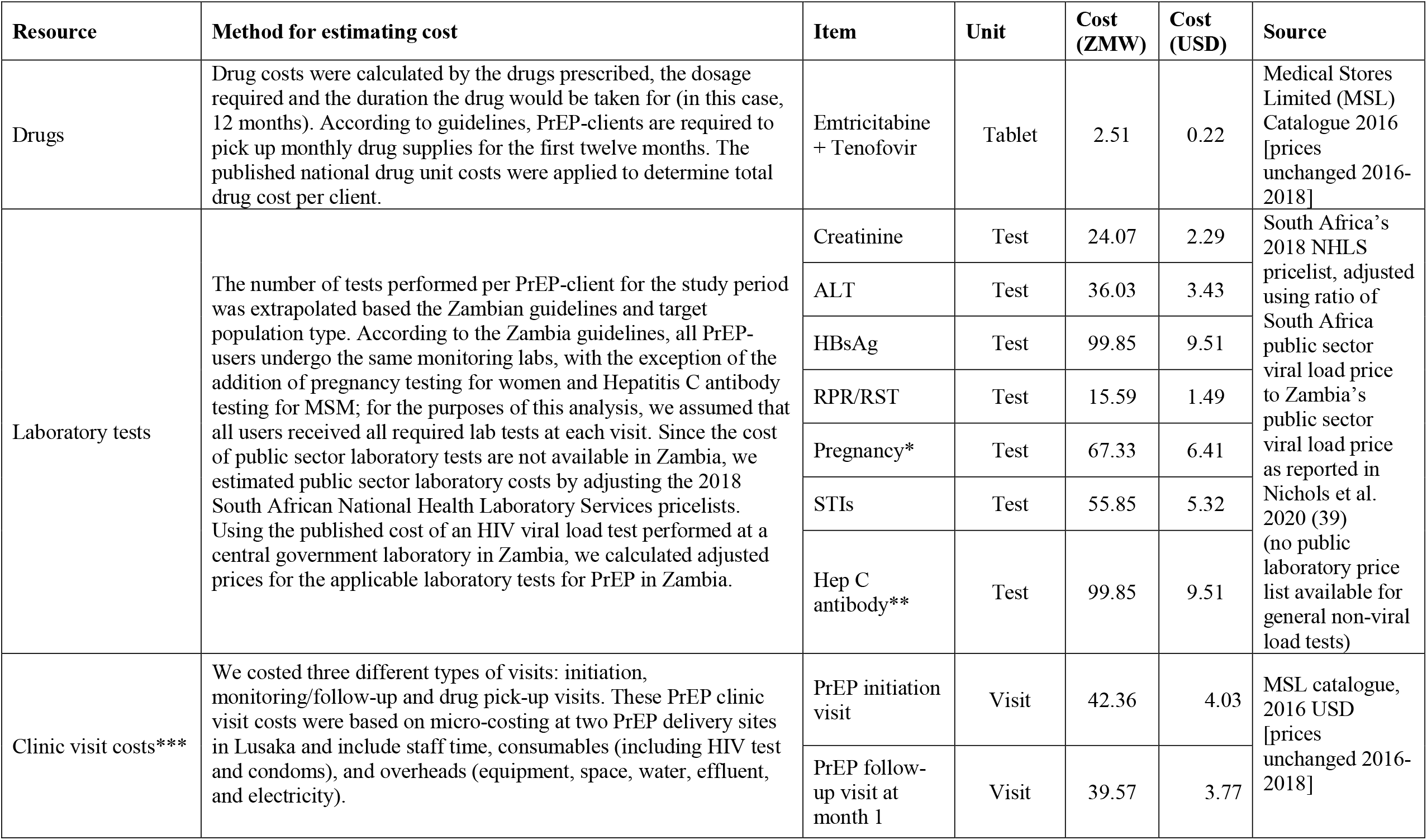
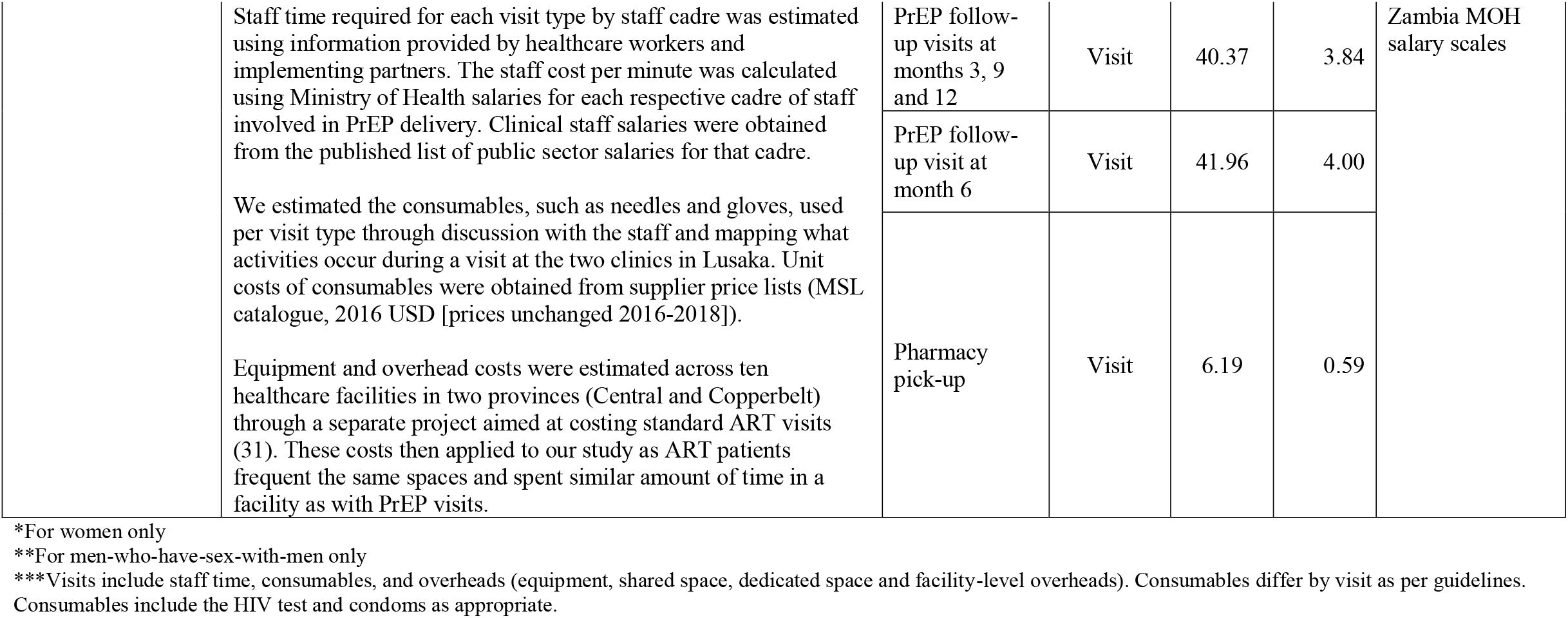
Cost methods and unit costs for direct service delivery costing.

| Resource | Method for estimating cost | Item | Unit | Cost (ZMW) | Cost (USD) | Source |
| --- | --- | --- | --- | --- | --- | --- |
| Drugs | Drug costs were calculated by the drugs prescribed, the dosage required and the duration the drug would be taken for (in this case, 12 months). According to guidelines, PrEP-clients are required to pick up monthly drug supplies for the first twelve months. The published national drug unit costs were applied to determine total drug cost per client. | Emtricitabine + Tenofovir | Tablet | 2.51 | 0.22 | Medical Stores Limited (MSL) Catalogue 2016 [prices unchanged 2016-2018] |
| Laboratory tests | The number of tests performed per PrEP-client for the study period was extrapolated based the Zambian guidelines and target population type. According to the Zambia guidelines, all PrEP-users undergo the same monitoring labs, with the exception of the addition of pregnancy testing for women and Hepatitis C antibody testing for MSM; for the purposes of this analysis, we assumed that all users received all required lab tests at each visit. Since the cost of public sector laboratory tests are not available in Zambia, we estimated public sector laboratory costs by adjusting the 2018 South African National Health Laboratory Services pricelists. Using the published cost of an HIV viral load test performed at a central government laboratory in Zambia, we calculated adjusted prices for the applicable laboratory tests for PrEP in Zambia. | Creatinine | Test | 24.07 | 2.29 | South Africa's 2018 NHLS pricelist, adjusted using ratio of South Africa public sector viral load price to Zambia's public sector viral load price as reported in Nichols et al. 2020 (39) (no public laboratory price list available for general non-viral load tests) |
|  |  | ALT | Test | 36.03 | 3.43 |  |
|  |  | HBsAg | Test | 99.85 | 9.51 |  |
|  |  | RPR/RST | Test | 15.59 | 1.49 |  |
|  |  | Pregnancy* | Test | 67.33 | 6.41 |  |
|  |  | STIs | Test | 55.85 | 5.32 |  |
|  |  | Hep C antibody** | Test | 99.85 | 9.51 |  |
| Clinic visit costs*** | We costed three different types of visits: initiation, monitoring/follow-up and drug pick-up visits. These PrEP clinic visit costs were based on micro-costing at two PrEP delivery sites in Lusaka and include staff time, consumables (including HIV test and condoms), and overheads (equipment, space, water, effluent, and electricity). | PrEP initiation visit | Visit | 42.36 | 4.03 | MSL catalogue, 2016 USD [prices unchanged 2016-2018] |
|  |  | PrEP follow-up visit at month 1 | Visit | 39.57 | 3.77 |  |

|  |  |  |  |  |  |
| --- | --- | --- | --- | --- | --- |
| Staff time required for each visit type by staff cadre was estimated using information provided by healthcare workers and implementing partners. The staff cost per minute was calculated using Ministry of Health salaries for each respective cadre of staff involved in PrEP delivery. Clinical staff salaries were obtained from the published list of public sector salaries for that cadre.<br>We estimated the consumables, such as needles and gloves, used per visit type through discussion with the staff and mapping what activities occur during a visit at the two clinics in Lusaka. Unit costs of consumables were obtained from supplier price lists (MSL catalogue, 2016 USD [prices unchanged 2016-2018]).<br>Equipment and overhead costs were estimated across ten healthcare facilities in two provinces (Central and Copperbelt) through a separate project aimed at costing standard ART visits (31). These costs then applied to our study as ART patients frequent the same spaces and spent similar amount of time in a facility as with PrEP visits. | PrEP follow-up visits at months 3, 9 and 12 | Visit | 40.37 | 3.84 | Zambia MOH salary scales |
|  | PrEP follow-up visit at month 6 | Visit | 41.96 | 4.00 |  |
|  | Pharmacy pick-up | Visit | 6.19 | 0.59 |  |

\*For women only
\*\*For men-who-have-sex-with-men only
\*\*\*Visits include staff time, consumables, and overheads (equipment, shared space, dedicated space and facility-level overheads). Consumables differ by visit as per guidelines. Consumables include the HIV test and condoms as appropriate.

Annualized program start-up, provider, and user support costs were based on program expenditure data and the number of PrEP sites and clients in 2019. Program start-up costs included training (annualized over two years), initial client demand creation activities and communication. Program support costs included recurring costs such as ongoing technical assistance and patient support costs (for example, peer navigators, peer support groups, social media support). We verified costing data via interviews with clinical staff and implementing partners and triangulation with program records. We analyzed costs in Excel 2018 (Microsoft, Redmond, USA). Ethical approval, as well as a consent process, were not required for this study as this was not human subjects’ research, given that only aggregate publicly available program data were used.

### Analysis

We estimated the annual cost of providing PrEP services assuming fidelity to the expanded 2018 Zambian PrEP guidelines for 12-months of PrEP (Table 1). We estimated patient resource use based on these guidelines from the day of PrEP initiation for 12 months, assuming continuous PrEP use for the duration. These direct service delivery costs included drugs, laboratory tests, and clinic visits. Assuming full adherence to guidelines, we then applied the unit costs to the resources expected to be used over the 12-month period by sex and risk group (Table 2).

Furthermore, we simulated a cohort of 1000 individuals taking PrEP over a 12-month period under two scenarios: 1) all PrEP-clients took PrEP continuously for 12-months and 2) using the actual continuation rates from programmatic PrEP data for visits at months 1, 3, 6, 9 and 12 for men, women and MSM. Using the population-specific service delivery costs, we determined a total cost per month based on these continuation rates. We calculated the cumulative number of protected months for each visit over a 12-month period and, using this, calculated the cost of PrEP per person-month effectively covered on PrEP.

## Results

The number of PrEP-clients and sites varied across programs, with the average number of PrEP-clients per site ranging from 3.6 in the Integrated 1 program and almost 70 in the AGYW program (Table 3). Start-up costs per site ranged from just over $400 for the Integrated 3 program to over $5000 for the AGYW program. However, this cost difference between the two programs disappeared when taking into account the number of PrEP-clients per site, with both programs costing $73 per PrEP-client per site. The Integrated 1 site incurred the largest start-up costs per PrEP-client per site at $230, while the Integrated 2 was the least costly at $26 per PrEP-client per site.

**Table 3:** 2018 costs of PrEP-provision according to Zambian guidelines (USD)

|  | Program Type |  |  |  |  |
| --- | --- | --- | --- | --- | --- |
|  | AGYW | FSW/MSM | Integrated 1 | Integrated 2 | Integrated 3 |
| **Number of PrEP clients initiated in program** | 908 | 213 | 322 | 214 | 148 |
| **Number of total sites** | 13 | 4 | 90 | 4 | 27 |
| **Average number of PrEP-clients/site** | 69.8 | 53.3 | 3.6 | 53.5 | 5.5 |
| **Start-up costs/site (2018)** |  |  |  |  |  |
| Community sensitization/consultation | 0 | 0 | 156 | 654 | 242 |
| Training | 3979 | 2844 | 668 | 308 | 160 |
| Site assessments | 0 | 0 | 0 | 422 | 0 |
| Equipment | 1149 | 1149 | 0 | 0 | 0 |
| **Average cost of start-up/site** | **5128** | **3992** | **824** | **1384** | **403** |
| **Average cost of start-up/site/PrEP-client** | **73** | **75** | **230** | **26** | **73** |
| **Recurrent costs** |  |  |  |  |  |
| **Programmatic support costs/PrEP-client** |  |  |  |  |  |
| Technical support | 0 | 0 | 0 | 154 | 265 |
| Training | 15 | 19 | 124 | 17 | 26 |
| PrEP user support | 63 | 83 | 0 | 0 | 85 |
| PrEP provider support | 13 | 0 | 193 | 0 | 0 |
| Demand creation communication | 22 | 31 | 4 | 0 | 0 |
| **Average programmatic support costs/PrEP-client (% of recurrent costs)** | **113 (35%)** | **133 (38%)** | **321 (61%)** | **171 (45%)** | **377 (64%)** |
| **Direct service delivery costs/PrEP-client** |  |  |  |  |  |
| Staff & Overheads | 46 | 46 | 46 | 46 | 46 |
| Labs and monitoring | 64 | 73 | 65 | 65 | 65 |
| Consumables | 13 | 13 | 13 | 13 | 13 |
| PrEP Drugs | 86 | 86 | 86 | 86 | 86 |
| **Average direct service delivery costs/PrEP-client (% of recurrent costs)** | **208 (65%)** | **217 (62%)** | **209 (39%)** | **209 (55%)** | **209 (36%)** |
| **Total recurrent cost/PrEP-client/year** | **320** | **350** | **530** | **381** | **586** |
| **Total average cost/PrEP-client/year** | **394** | **425** | **760** | **406** | **659** |
| **Total cost/person-month\*** | **33** | **35** | **63** | **34** | **55** |

Recurrent costs per PrEP-client also varied by program, driven by the differences in programmatic support costs. These ranged from $113 for the AGYW program to $377 for the third integrated program. Direct service delivery costs did not vary greatly, with the variation due to the different monitoring laboratory tests guidelines require for each target population. The proportion of recurrent costs attributable to programmatic support costs ranged from just over a third (35%) for the AGYW-focused program to almost two-thirds for the third integrated program (64%), with the two priority population focused programs spending a lower proportion of these costs on programmatic support costs than the integrated programs (Figure 1). The drug costs carried the highest proportion of the service delivery costs at about 40%, followed by monitoring labs at about a third of the cost. However, when compared to the costs of drugs and labs as a proportion of the entire recurrent program costs, this fell to between 15-27% and 11-21% respectively.

**Figure 1:**
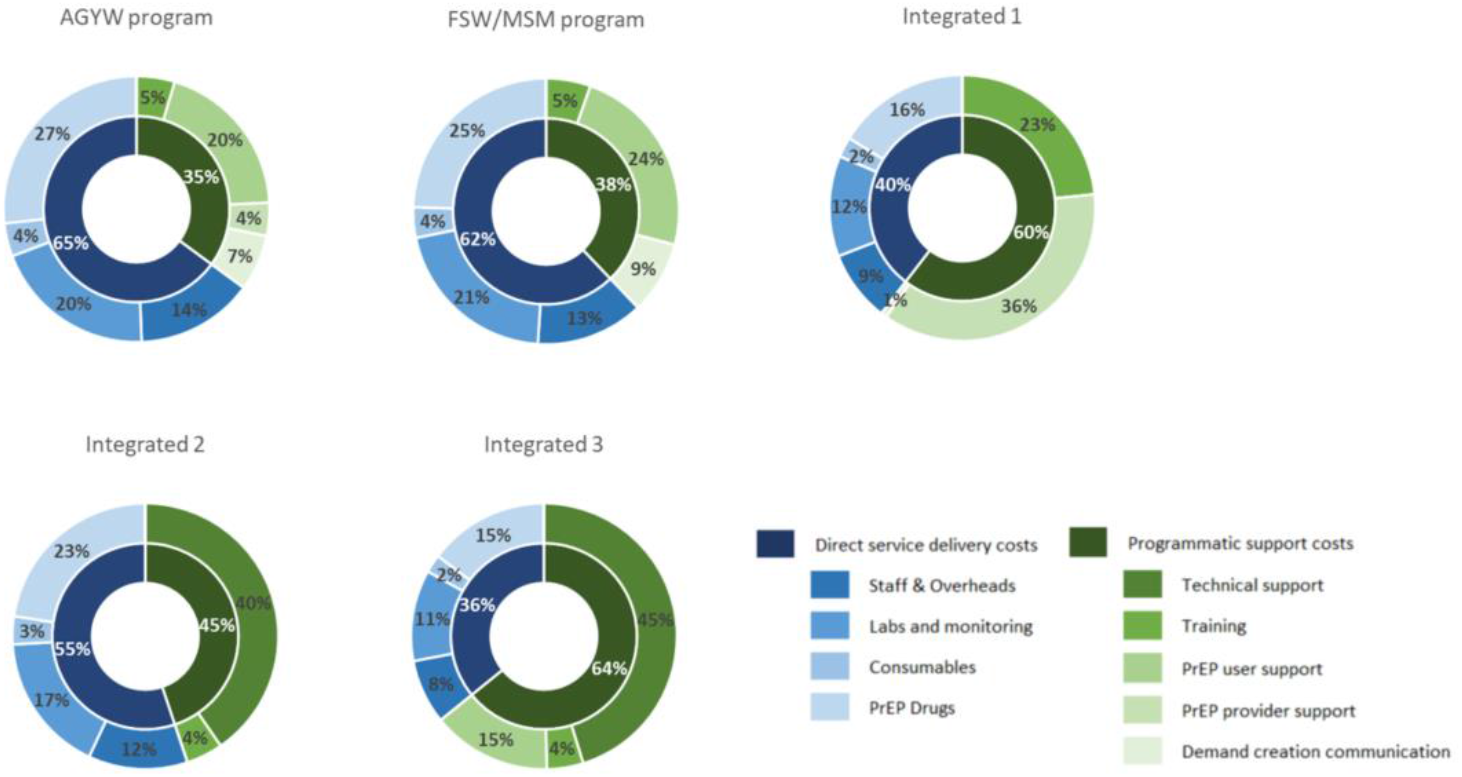
Proportion of total annual cost per PrEP-client by cost inputs.

When taking the average number of PrEP-clients initiated per site into account, and combining program start-up and recurrent support costs and assuming 12 months of continuous PrEP-use, the annual cost per PrEP initiate in 2018 varied greatly by program type, with two of the integrated programs being most costly at $760 and $660 per PrEP-initiate. The FSW/MSM-targeted program and third integrated program cost $425 and $406 per PrEP initiate respectively. The least costly program per PrEP initiate, considering start-up costs, number of sites and site volume was the AGYW program at $394 per PrEP initiate. These cost differences were driven largely by the differences in the average number of PrEP-clients initiated per site across each program. The total cost per PrEP-client per person-month, assuming 12 months of continuous PrEP use was $63, almost double that of the AGYW-focused program at $33.

Comparing two simulation scenarios, one with perfect PrEP persistence and the other using PrEP continuation data from Zambia, we see that the cost of PrEP provision per person-month effectively covered on PrEP decreases over time for all groups under both scenarios (Figure 2). However, this decrease is less substantial when there is imperfect persistence. For example, the cost of PrEP provision per person month among MSM is 76% greater at 12 months when using actual data on persistence in that risk group as compared to assuming perfect persistence. For women, this difference is smaller at 27% ($20 per person at 12 months with actual persistence rates versus $16 for perfect persistence). The smallest difference is among men with an 18% greater cost of PrEP provision per person month at 12 months.

**Figure 2:**
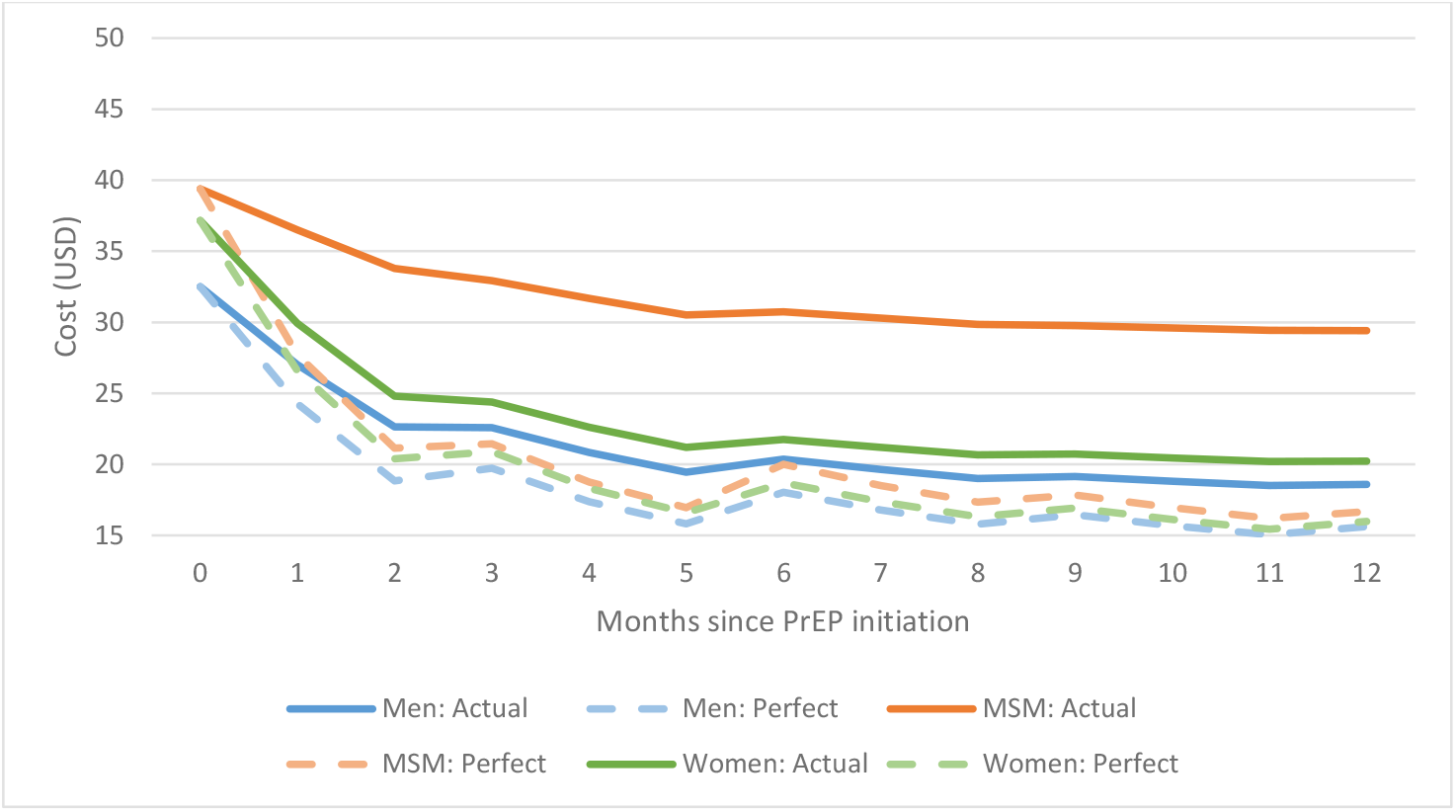
Cost of PrEP provision per person-month effectively covered on PrEP, using aggregate persistence data from partner data (USD) (Service delivery costs only)

## Discussion

Modelling studies for PrEP in SSA indicate that PrEP can be cost effective when targeted to specific high-risk groups (40); however, most models have not used data from real-world implementation settings. To date only a few studies have reported the cost of providing PrEP to priority populations in SSA using data from demonstration projects or routine implementation. In this study, we address this gap and add to the small but growing PrEP cost literature by describing the guideline-based costs of PrEP service delivery in the context of a generalized epidemic in an LMIC. We estimated that the overall average cost to provide PrEP to an at-risk individual for a full 12 months in Zambia ranged from $394 to $760, or $33 to $63 per person-month on PrEP. This varied by the different service delivery models, with programs focused on reaching priority populations (AGYW, FSW/MSM) generally costing less than those integrated into existing healthcare facilities, when taking into consideration the average number of PrEP-clients per site. Our analysis shows that recurrent PrEP-provision costs are variable depending on the type of program; however, we estimate that the costs are between $320 and $586 per PrEP initiate depending on the program type. These costs are based solely on the Zambian PrEP policy, which assumes that all patients were treated precisely according to these guidelines, with no variation among patients based on condition, logistical challenges, or other potential variants.

The costs of PrEP provision we report in this study, ranging from $394 to $760 are generally higher than those documented in other costing studies from the region. In Kenya and Uganda, the incremental cost of providing PrEP to serodiscordant couples under study settings was just over $305 and $408 respectively (25, 27), while in South Africa PrEP provision cost about $127 per year for FSW (24). A study in Zimbabwe reported the cost per PrEP client initiated as $238 while another study in Kenya reported an overall cost of $256 (28, 41). Compared to our calculated costs per person-month, ranging from $33 to $63, other studies were also lower at $26.52 under study settings or $16.54 under MOH scenario and $21.32 and $28.92 study, $14.52 for adolescent girls and young women (29, 42, 43). One possible reason for these differing results are the relatively low PrEP client volumes in these programs compared to those reported in other studies, which ranged from 219 to almost 5000. In Kenya, authors report that the cost per client-month of PrEP dispensed is reduced substantially if PrEP delivery is scaled up. This is supported in other HIV scale-up studies that have demonstrated that early implementation costs are greater per patient served (“U-shaped incremental cost curve”) (44-46). Understanding where we are on the U-shaped curve will assist with planning and to ensure programs are adequately resourced. Similar to HIV treatment programs focusing on hard-to-reach key populations, targeted PrEP programs require substantial investment in order to establish and maintain successful programs (47-50). More intensive programmatic inputs and larger PrEP-user volumes are required per site for the key population-focused programs to reach the minimum cost/user; however, this investment may be necessary to effectively reach these groups, especially as more than half of all new HIV infections were among key populations in 2018, despite being a minority of the overall population (51).

It should also be noted that the start-up cost of scaling up any of these programs nationally likely exceeds the simple multiplication of start-up costs and number of sites. Particularly for the integrated approach, many of the sites that have been reached to date have been relatively easy to reach. The cost to scale-up to all facilities may therefore start to increase once smaller health facilities that serve fewer patients are reached (the other side of the U-shaped incremental cost curve), with additional travel and staff costs to get these sites started (52). There are now more than 1500 health facilities in Zambia that provide ART services (e.g. these could potentially also all provide PrEP services), of which 24% provide service to 80% of patients, so an effort to scale-up PrEP to all facilities may be cost-prohibitive.

Using PrEP persistence data from Zambia to simulate the cost of PrEP provision over time, we observe that low persistence results in higher costs per PrEP-client, up to 76%. Therefore, understanding PrEP persistence in the population is key to planning and budgeting. Our data show that it is costly to initiate someone on PrEP who only persists on PrEP for a few months. This does not necessarily mean that a PrEP program is not cost-effective; it just means that this low-usage, or in-and-out cycling, needs to be accounted for when planning PrEP provision and program scale-up. These costs vary with the duration of at-risk periods when PrEP is required – longer periods on PrEP require more visits and medications resulting in high costs of continual PrEP use per year per PrEP-client, though lower per month costs. Preliminary research has shown that, despite heterogeneity within sub-groups of PrEP clients, PrEP persistence in Zambia is low, with retention at three months only 27% in one study (33, 34). This means that the cost per initiate will likely decline rapidly for a given PrEP initiation cohort. Furthermore, the cost per PrEP initiate may be lower depending on how many carry-over PrEP clients come from 2018 into 2019.

This analysis is subject to a number of limitations. First, we relied on implementing partners and provider reports and some resource use may not have been recorded. We were unable to directly observe time spent for PrEP alone from staff time on other primary care and relied on provider recall and estimates to determine time per visit. This may mean our staff cost estimates are lower than would be expected in a low HIV prevalence setting or where care is not integrated. We also excluded salary costs of ancillary implementing partner staff who were involved in PrEP-program planning as they were reported inconsistently across partners. Including these costs would further increase this cost per PrEP client served. These lowest cost thresholds will not apply to truly integrated models where capacity and resources are shared. Furthermore, this analysis does not investigate the effect that changes in integration and standardization would have on the cost of PrEP provision across different clinic sizes. This is an important factor when thinking about PrEP provision to small or more rural clinics where high PrEP-user volumes may not be achievable. Ensuring access, however, even in remote or low-volume facilities is important to ensure equitable PrEP access, and costs associated with a lower PrEP client volume should be planned for. As this analysis is based on national guidelines and does not include patient or program outcomes, we are not making any conclusions comparing the effectiveness of the various models, but are providing budgetary information only. Further costing work is needed to elucidate the effect of PrEP persistence on the overall costs of PrEP provision. Despite these limitations, this study provides a robust estimate of the costs of PrEP provision under three different service delivery models intended to guide future scale-up. This information will support budgeting and financial planning for PrEP services as Zambia scales up HIV prevention and access to services to achieve national and international targets.

## Conclusions

Given finite resources, methods to ensure low incremental costs for PrEP provision are important. This work demonstrates that the routine implementation and service delivery of PrEP may be more costly than modelling and early demonstration projects indicated. In addition to direct delivery cost, PrEP programs incur substantial costs for demand creation and PrEP-user support. The costs involved in these activities to reach key and priority populations should not be discounted when budgeting for PrEP program scale-up.

## Data Availability

All data produced in the present study are available upon reasonable request to the authors

## Competing interests

CH, LL, and BEN report grants from United States Agency for International Development (USAID), during the conduct of the study. All other authors have no competing interests.

## Authors’ contributions

CH and BEN were responsible for the initial concept for the paper

LCL, CvR, HO, CWC, MN, CM, LM, CAR were responsible for critical input into development of the paper

CH and BEN wrote the first draft of the manuscript

LCL, CvR, HO, CWC, MN, CM, LM, CAR contributed to the writing of the manuscript

All authors reviewed and approved the final draft of the manuscript

## Acknowledgements

USAID funded this work through cooperative agreement AID-OAA-A-15-00070 (to CH, LCL and BEN). Research reported in this publication was also supported by the Fogarty International Center and National Institute of Mental Health through the National Institutes of Health award number D43 TW010543 (to CH) and K01MH119923 (LCL). The funders had no role in study design, data collection and analysis, decision to publish, or preparation of the manuscript. The authors’ views expressed in this publication do not necessarily reflect the views of USAID or the US Government.

## Notes

### Competing Interest Statement

The authors have declared no competing interest.

### Author Declarations

Source data are publicly available from a manuscript published on BMJ Open: https://pubmed.ncbi.nlm.nih.gov/34244265/

